# Patterns of impaired neurocognitive performance on Global Neuropsychological Assessment (GNA), and their brain structural correlates in recent-onset and chronic schizophrenia: A pilot study

**DOI:** 10.1101/2022.04.12.22273462

**Authors:** Vineeth Mohan, Pravesh Parekh, Ammu Lukose, Sydney Moirangthem, Jitender Saini, David J Schretlen, John P John

## Abstract

Cognitive deficits are established as a fundamental feature of schizophrenia; however, their pattern and how they are affected by chronicity are still unclear. Although a generalized stable impairment affecting multiple cognitive domains is commonly seen from the onset, some longitudinal studies have shown evidence of neuroprogression, and selective deterioration in certain cognitive domains. We assessed cognitive performance in patients with recent-onset (*n* = 17, duration of illness ≤ 2 years) and chronic schizophrenia (*n* = 14, duration ≥ 15 years), and healthy adults (*n* = 16) using the Global Neuropsychological Assessment and examined correlations between cognitive scores and gray matter volumes computed from T1-weighted MRI images. We also measured and analyzed differences between patient groups for negative and positive symptoms, psychotic exacerbations, and medication exposure, and studied their correlations with cognitive performances. We observed cognitive deficits affecting multiple domains in both recent-onset and chronic schizophrenia samples. Selectively greater impairment of perceptual comparison/processing speed was found in adults with chronic schizophrenia (*p* = 0.009, η^2^_partial_ = 0.25). In the full sample (*n* = 47), perceptual comparison speed correlated significantly with gray matter volumes in the anterior and medial temporal lobes, predominantly on the left side (TFCE, FWE *p* < 0.01). These results indicate that along with generalized deficit across multiple cognitive domains, selectively greater impairment of perceptual comparison/processing speed appears to characterize chronic schizophrenia. This pattern might indicate an accelerated or premature cognitive aging. Gray matter volumetric deficits in the anterior-medial temporal lobes especially of left side might underlie the impaired perceptual comparison/processing speed seen in schizophrenia.

## 1. Introduction

Neurocognitive deficits are considered endophenotypes of schizophrenia (Greenwood et al., 2019). The pattern and magnitude of cognitive impairment in schizophrenia vary widely across studies (Fioravanti et al., 2012), probably due to differences in the tools used, the illness phases when assessments were done, and phenotypic heterogeneity in schizophrenia (Wolfers et al., 2018). A generalized deficit spanning multiple domains is the most replicated finding, with relatively large effect sizes for verbal declarative memory, working memory and processing speed (Donati et al., 2020).

Cognitive deficits of schizophrenia may manifest even before the onset of overt symptoms, in the at-risk state or the prodrome (Addington and Lewis, 2010; Fusar-Poli et al., 2012; Seidman et al., 2016). Cross-sectional studies in different illness states such as neuroleptic-naïve, recent-onset, and chronic schizophrenia have reported comparable magnitudes of the overall cognitive impairment (Carrión et al., 2018). However, differentially severe deficits in certain domains like working memory, verbal fluency, executive functioning, and processing speed have also been reported in chronic schizophrenia compared to recent-onset and prodromal/high risk individuals (Liu et al., 2019). Longitudinal studies have not observed substantial worsening of cognitive deficits with increasing duration of illness (Heilbronner et al., 2016); however, there is evidence of more pronounced decline in executive functioning, verbal fluency, and visuomotor processing than what is seen in normal aging (Rund et al., 2016; Valsdottir et al., 2020; Zanelli et al., 2019). Additionally, older patients can show greater cognitive dysfunction than younger patients relative to age-matched healthy individuals (Loewenstein et al., 2012; Mosiołek et al., 2016; Murante and Cohen, 2017). Thus, the effect of chronicity on cognitive dysfunction in schizophrenia might be influenced by aging in specific cognitive domains (Lee et al., 2020). However, few studies have examined the effect of chronicity on cognition over a period of 10 years or more (Albus et al., 2020; Fett et al., 2020).

Most of the test batteries that have been used to assess cognitive functioning in schizophrenia take at least two hours to complete (Harvey, 2013; Keefe, 2004; Pietrzak et al., 2009). A common problem with long neuropsychological batteries is fatigue which can potentially affect test performance. A general psychomotor slowness may be seen in patients with schizophrenia due to medication effects, sleep impairment, primary negative symptoms etc. that could make patients more prone to get fatigued (Irani et al., 2012). Hence a brief battery that can cover the relevant cognitive domains would be beneficial to study the profile of cognitive impairment in schizophrenia.

The Global Neuropsychological Assessment (GNA) is a brief neuropsychological battery developed as part of the initiative to develop global norms for cognitive tests by a public non-profit organization (“Global Neuropsychology, Inc. GNI,” 2018). It is available in five alternate forms. The GNA consists of seven main tests — Immediate Story Memory Trial 1 and 2, Perceptual Comparison, Digit Span Forward and Backward, Delayed Story Memory, Verbal Fluency and Category Switching, Spatial Span Forward and Backward, and Patient Health Questionnaire-4 (PHQ-4). These tests are adaptations or modifications of extensively studied and validated cognitive tests, and the battery is currently under the process of validation worldwide. The GNA can be administered in less than 25 (healthy adults) to 30 minutes (patients) in most cases. Preliminary validation and test-retest reliability of this battery in mild cognitive impairment and Alzheimer’s disease were recently reported (Olson et al., 2021), as were analyses of GNA interform differences and the effects of repeated administration (Smerbeck et al., 2021). The utility of this battery for detecting neurocognitive impairment in schizophrenia has not been examined so far.

In this pilot study, we aimed to examine the utility of the GNA for detecting neurocognitive abnormalities in schizophrenia, determine whether these impairments are differentially present in recent onset vs. chronic schizophrenia, and correlate neurocognitive functioning with brain morphometry in these samples.

## 2. Materials and Methods

### 2.1 Participants

This cross-sectional study was carried out on patients with recent-onset schizophrenia (ROSZ) with duration of illness 2 years or less (*n* = 17), chronic schizophrenia (CHSZ) with duration of illness 15 years or more (up to 25 years) (*n* = 14), and healthy comparison subjects (HCS) (*n* = 16). Patients who met the criteria as per the Diagnostic and Statistical Manual of Mental Disorders (5^th^ ed.; DSM-5; American Psychiatric Association, 2013) for schizophrenia based on the Structured Clinical Interview for DSM-5 Disorders—Clinician version (SCID-5-CV) (First et al., 2016) were recruited from the outpatient and inpatient sections of the Department of Psychiatry, NIMHANS, Bangalore during the period from May 2019 – February 2020. Healthy subjects were recruited by word of mouth from hospital staff and students at NIMHANS, during the same period. The study was approved by the NIMHANS Institutional Ethics Committee [NO.NIMH/DO/IEC (BEH.Sc.DIV)/2019, dated 25^th^ May 2019], and research participants were recruited after obtaining their written informed consent. Healthy subjects were screened to rule out psychiatric or other medical disorders, substance use disorders, and first-degree family history of psychiatric disorders, using a study-specific checklist. Patients with comorbid psychiatric disorders including substance use disorders (except tobacco use disorder), and other medical/neurological disorders were excluded from the study. Patients who have had electroconvulsive therapy (ECT) within the previous 6 months, or who had been on regular dose of benzodiazepines over the previous 2 weeks or having received SOS treatment within the previous 48 hours were also excluded. Handedness was assessed using Edinburgh Handedness Inventory (Oldfield, 1971) and only right-handed subjects were recruited. All the participants underwent the clinical and neurocognitive assessments as well as MR imaging either on the same day or within 1-2 days of each other.

### 2.2 Socio-demographic and clinical details of the study samples

Clinical symptom ratings were done for all the patients using the Psychotic Symptom Rating Scales (PSYRATS) (Haddock et al., 1999), the Scale for the Assessment of Negative Symptoms (SANS) (Andreasen, 1983), and the Scale for the Assessment of Positive Symptoms (SAPS) (Andreasen, 1984). Medication associated abnormal movements were measured using the Abnormal Involuntary Movement Scale (AIMS) (Guy, 1976), the Simpson-Angus Scale (SAS) (Simpson and Angus, 1970), and the Barnes Akathisia Rating Scale (BARS) (Barnes, 1989) (see **Table 2**). The socio-demographic details of the study participants are summarized in **Table 1**.

**Table 1.**
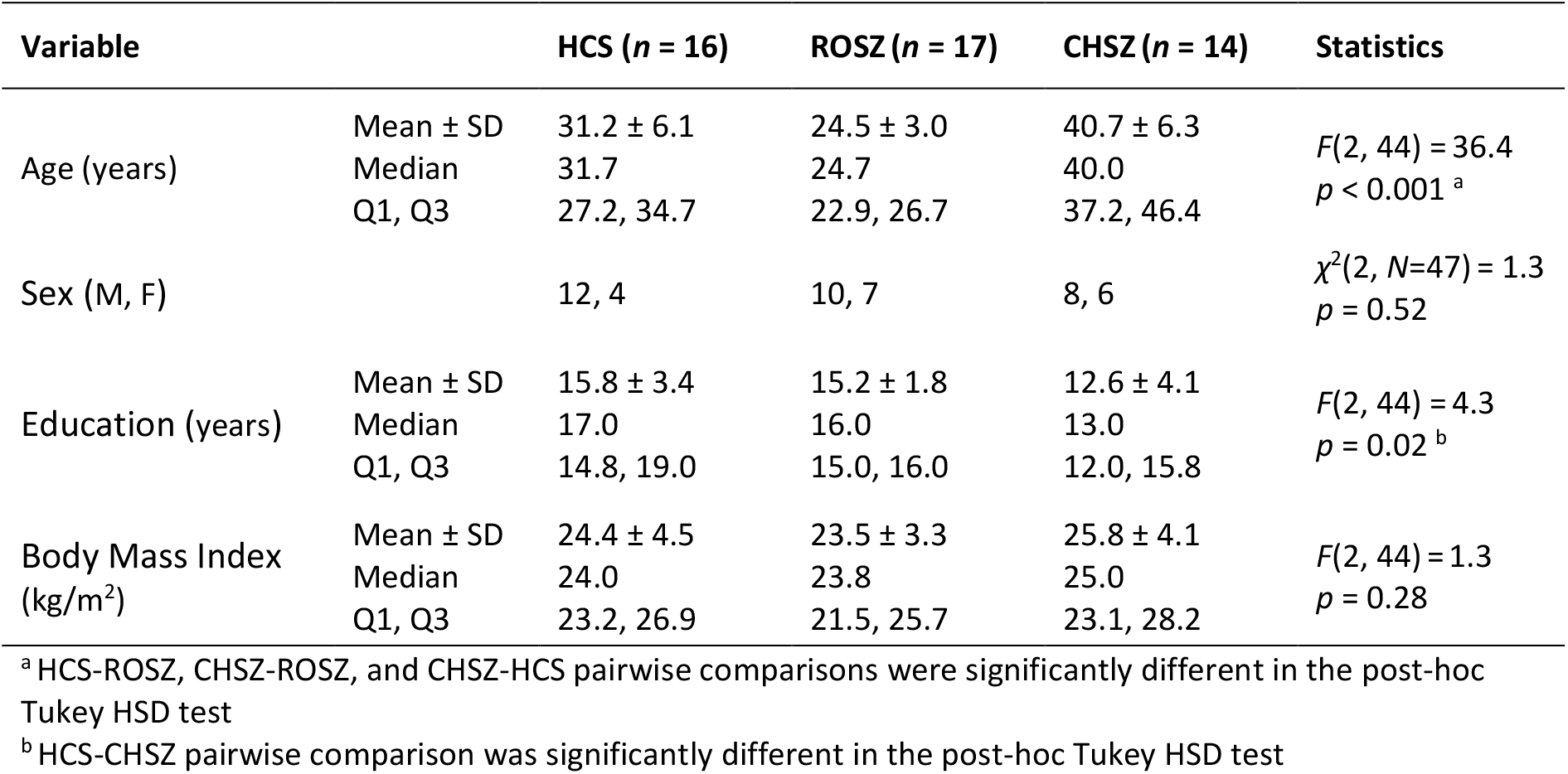
Socio-demographic details of the study samples

**Table 2.** Clinical details of the schizophrenia samples

| Variable | ROSZ (n = 17)<br>Mean ± SD | CHSZ (n = 14)<br>Mean ± SD | Welch's t-test, t(df), p-value |
| --- | --- | --- | --- |
| Age at onset | 22.8 ± 3.3 | 21.8 ± 6.0 | $t(19.2) = 0.58, p = 0.57$ |
| Psychotic Symptom Rating Scales (PSYRATS) | 30.8 ± 18.9 | 18.3 ± 17.0 | $t(28.7) = 1.93, p = 0.06$ |
| Scale for the Assessment of Negative Symptoms (SANS) | 35.6 ± 17.4 | 33.9 ± 21.4 | $t(25.0) = 0.24, p = 0.81$ |
| Scale for the Assessment of Positive Symptoms (SAPS) | 28.4 ± 16.3 | 18.5 ± 18.3 | $t(26.4) = 1.58, p = 0.12$ |
| Abnormal Involuntary Movements Scale (AIMS) | 0.5 ± 1.6 | 0.5 ± 1.0 | $t(27.5) = 0.06, p = 0.95$ |
| Simpson Angus Scale (SAS) | 1.8 ± 2.1 | 1.9 ± 0.9 | $t(27.5) = -0.18, p = 0.86$ |
| Barnes Akathisia Rating Scale (BARS) | 0.3 ± 0.9 | 0.4 ± 0.8 | $t(28.9) = -0.23, p = 0.86$ |
| Current antipsychotic dose in risperidone equivalents (mg) | 5.3 ± 2.4 | 8.0 ± 4.8 | $t(18.2) = -1.94, p = 0.07$ |
| Current anticholinergic (trihexyphenidyl) dose (mg) | 2.6 ± 1.8 | 2.4 ± 1.6 | $t(28.9) = 0.26, p = 0.80$ |
| Cumulative antipsychotic exposure in dose-years of risperidone equivalents | 3.3 ± 2.6 | 58.5 ± 33.2 | $t(13.1) = -6.20, p < 0.001$ |
| Average daily dose of antipsychotic exposure in risperidone equivalents | 4.7 ± 1.9 | 5.8 ± 2.2 | $t(25.7) = -1.41, p = 0.17$ |
| Cumulative FGA exposure in dose-years of risperidone equivalents | 0.03 ± 0.14 | 7.0 ± 9.8 | $t(13.0) = -2.67, p = 0.02$ |

FGA, First Generation Antipsychotics

The risperidone equivalents of antipsychotics were calculated based on the defined daily doses (DDD) by the World Health Organization (WHO) Collaborating Centre for Drug Statistics Methodology (https://www.whocc.no/atc_ddd_index/) implemented in an excel conversion sheet by Leucht et al (http://www.cfdm.de/indexab2e.html?option=com_content&task=view&id=15&Itemid=29) (Leucht et al., 2016). In the absence of a gold standard method to compare antipsychotic exposures (Patel et al., 2013), this conversion sheet lists almost all the old and new antipsychotics based on a widely accepted and reliable method (Leucht et al., 2019).

### 2.3 Global Neuropsychological Assessment (GNA)

GNA covers most of the cognitive domains that have been reported in the literature to be impaired in schizophrenia (McCleery and Nuechterlein, 2019), namely, attention, verbal and visuo-spatial working memory, perceptual comparison/processing speed, verbal declarative memory (immediate and delayed story recall), and verbal fluency (category fluency and switching) (see **Table 3**).

**Table 3.** Tests in GNA and the cognitive domains assessed

| Test item in GNA | Cognitive domain |
| --- | --- |
| Immediate story memory—trial 1 and 2, and<br>Delayed story memory | Verbal declarative Memory |
| Perceptual comparison | Perceptual comparison/processing<br>speed |
| Digit span forward | Simple attention |
| Digit span backward | Verbal working memory |
| Verbal fluency—Category fluency (animal<br>names); Category switching (body parts/foods) | Verbal fluency/executive functioning |
| Spatial span forward and backward | Visuospatial working memory |

**Table 4.**
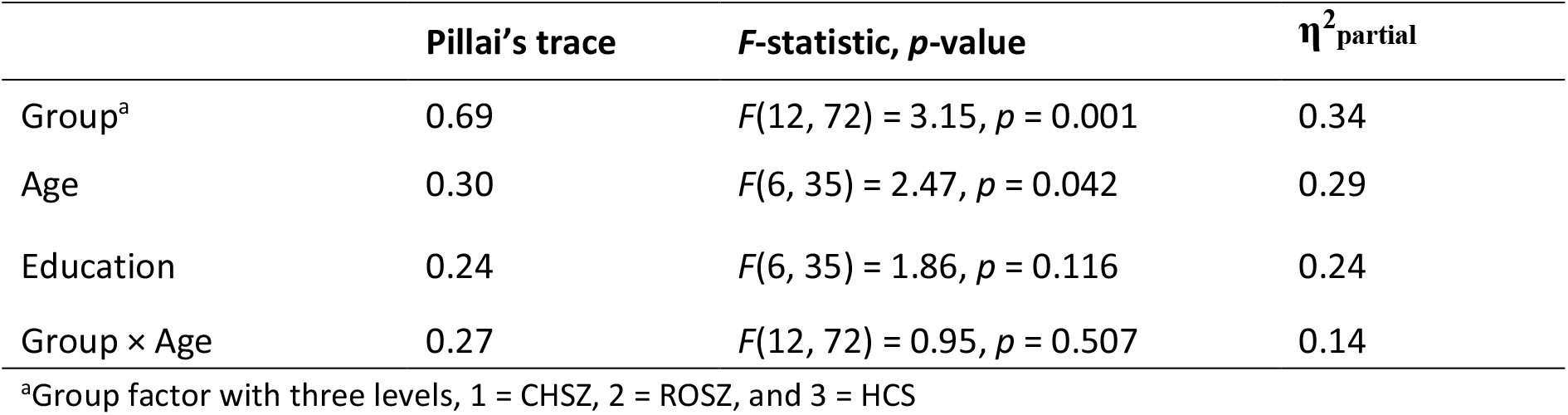
Multivariate GLM (MANCOVA) results (*n* = 47)

| | Pillai's trace | F-statistic, $p$-value | $\eta^2_{\text{partial}}$ |
| --- | --- | --- | --- |
| Group<sup>a</sup> | 0.69 | $F(12, 72) = 3.15, p = 0.001$ | 0.34 |
| Age | 0.30 | $F(6, 35) = 2.47, p = 0.042$ | 0.29 |
| Education | 0.24 | $F(6, 35) = 1.86, p = 0.116$ | 0.24 |
| Group × Age | 0.27 | $F(12, 72) = 0.95, p = 0.507$ | 0.14 |

<sup>a</sup>Group factor with three levels, 1 = CHSZ, 2 = ROSZ, and 3 = HCS

English, Hindi, and Malayalam versions of the GNA were administered according to the language preferences of the study participants. Bi-/multi-lingual mental health professional volunteers under the supervision of AL, VM, and JPJ ensured equivalence of the content of items and their intended meaning in the Hindi and Malayalam translations with those of the original English version (semantic and content equivalence).

The GNA battery was administered in single session, by a psychiatrist (VM). The tests were administered in the same sequence and as per the standard instructions for administration of the GNA. All the participants completed the full battery including all the tests. The average duration of the GNA session was 21.2 minutes in the healthy subjects and 25.8 minutes in the schizophrenia sample. The individual test items were scored and entered in a spreadsheet and visually double-checked for data entry errors before the analysis.

We converted the raw individual test scores to *z*-scores using the overall mean and pooled standard deviation of the three groups. Then, we grouped the *z*-scores into the six cognitive domains (see **Table 3**) by averaging the *z*-scores of the tests under each domain. Thus, the GNA test battery which had seven cognitive tests with 13 scorable items was reduced into six meaningful and comparable domains, which were then analyzed using R, version 4.0.5 (RStudio 1.3.1073) (RStudio Team, 2020).

### 2.4 Imaging methods (acquisition and pre-processing)

The MR images were acquired on a 3 Tesla Philips Ingenia CX scanner using a 32-channel head coil (TR 6.5 ms, TE 2.9 ms, flip angle 9°, 192 slices in sagittal orientation, voxel size 1mm isotropic). The T1-weighted images were checked for MR artefacts, motion, and structural abnormalities using a systematic quality check (QC) pipeline (Parekh et al., 2021). The scans were also reviewed independently by a neuroradiologist to rule out any gross morphological abnormality. We first set the origin of T1-weighted images approximately at the anterior commissure using acpcdetect v2.0 (Ardekani, 2022; Ardekani et al., 1997; Ardekani and Bachman, 2009) (https://www.nitrc.org/projects/art). The T1-weighted images were then segmented into gray matter, white matter and, cerebrospinal fluid tissue classes using the Computational Anatomy Toolbox (Gaser and Dahnke, 2016) (CAT, version 1727, http://www.neuro.uni-jena.de/cat/) with SPM12 (version 7771, http://www.fil.ion.ucl.ac.uk/spm) in the background running on MATLAB R2016a (MathWorks, Natick, Massachusetts, USA; https://www.mathworks.com). We then smoothed the resulting modulated normalized gray matter images by a Gaussian kernel of 6 mm full width at half maximum which were then used for voxel-level correlation of gray matter with cognitive scores (See 2.5.2).

### 2.5 Statistical Analysis

#### 2.5.1 Analysis of the neuropsychological data

We visualized the distribution of *z*-scores in each group using violin plots. Then, we calculated the Pearson’s correlations between cognitive domains. Additionally, to qualitatively examine the cognitive profiles of each group, we plotted the mean *z*-scores and their standard errors for each domain.

To quantify the between-group differences in ROSZ and CHSZ groups (with respect to the HCS group), we calculated the effect size for each cognitive domain (Hedge’s *g*), assuming unequal variance in scores between groups. Hedge’s *g* (Hedges, 1981) corrects for the bias in Cohen’s *d* (Cohen, 1988), given the small sample sizes in the present study. We used the package ‘effectsize’ (Ben-Shachar et al., 2020) in R for this calculation, which implements the method recommended by Delacre et al for groups with unequal variance (Delacre et al., 2021).

We performed a multivariate general linear model (GLM-MANCOVA) analysis with the six cognitive domain scores as dependent variables and group (factor with three levels - HCS, ROSZ and CHSZ), age, education, and group-by-age interaction as independent variables. We additionally did post-hoc univariate GLM (ANCOVA) analyses, to explore the cognitive domains that contributed the most to the group difference.

#### 2.5.2 Analysis of the neuroimaging data

To examine the association of cognitive scores with gray matter volumes, we performed a voxel-level correlation with cognitive scores in the overall sample (*n* = 47) and included the total intracranial volume (TIV) and age as covariates of no interest. We employed non-parametric statistical inference using the Threshold Free Cluster Enhancement (TFCE) algorithm implemented in the TFCE toolbox (version 210, http://www.neuro.uni-jena.de/tfce/). TFCE method combines the advantages of both voxel and cluster level inference (Smith and Nichols, 2009). We used the default settings (Smith method; 5000 permutations, cluster size weighting of *E* = 0.5) and examined the results at a stringent family-wise error rate threshold of FWER < 0.0083, to account for probable false positives due to the multiple tests done for correlating each of the six cognitive domains with gray matter volume (i.e., Bonferroni correction for six cognitive domains: *α*=0.05/6=0.0083).

## 3. Results

All the six cognitive domains showed significant correlations with each other, with the largest correlation being *r* = 0.72, between simple attention (digit span forward) and verbal working memory (digit span backward). All the pairwise correlations survived Bonferroni correction for multiple comparisons (*α*=0.05/15=0.0033) (see **Figure S2** in supplementary material). The mean and median cognitive performance *Z-*scores showed a pattern of HCS > ROSZ > CHSZ in all the domains (see **Figure 1** and **Figure *2***).

**Figure 1.**
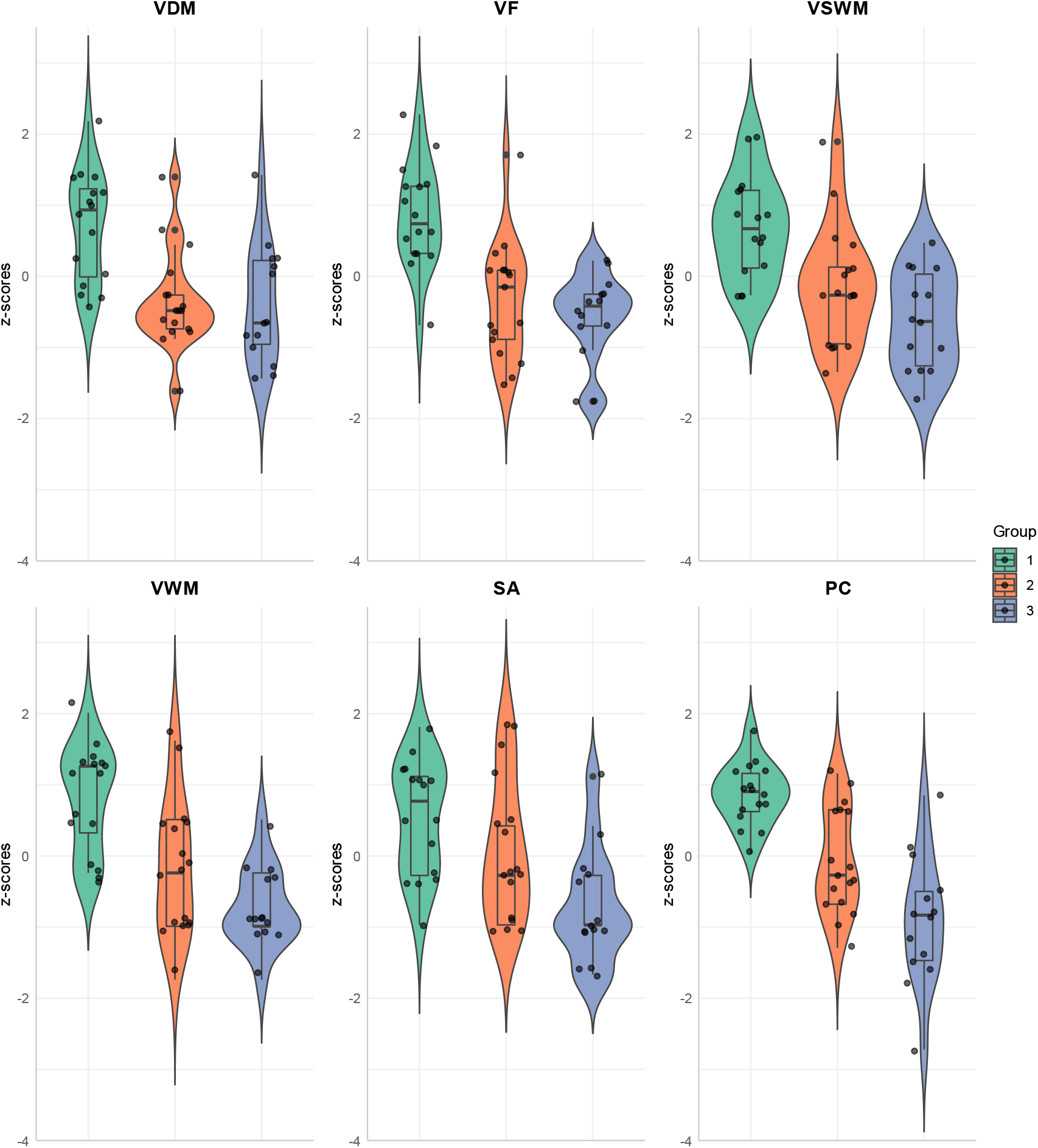
Violin plots depicting the domain-wise cognitive *z*-score distribution in the three groups. VDM, Verbal declarative memory; PC, Perceptual comparison; SA, Simple attention; VWM, Verbal working memory; VF, Verbal fluency; VSWM, Visuospatial working memory. As the violin plots are based on kernel density estimates, they extend beyond the range of actual scores in the sample depicted by the jittered dots. The solid horizontal lines within the boxplots represent the median *z*-score. Group 1: Healthy control subjects (HCS); Group 2: Recent-onset schizophrenia (ROSZ); Group 3: Chronic schizophrenia (CHSZ).

**Figure 2.**
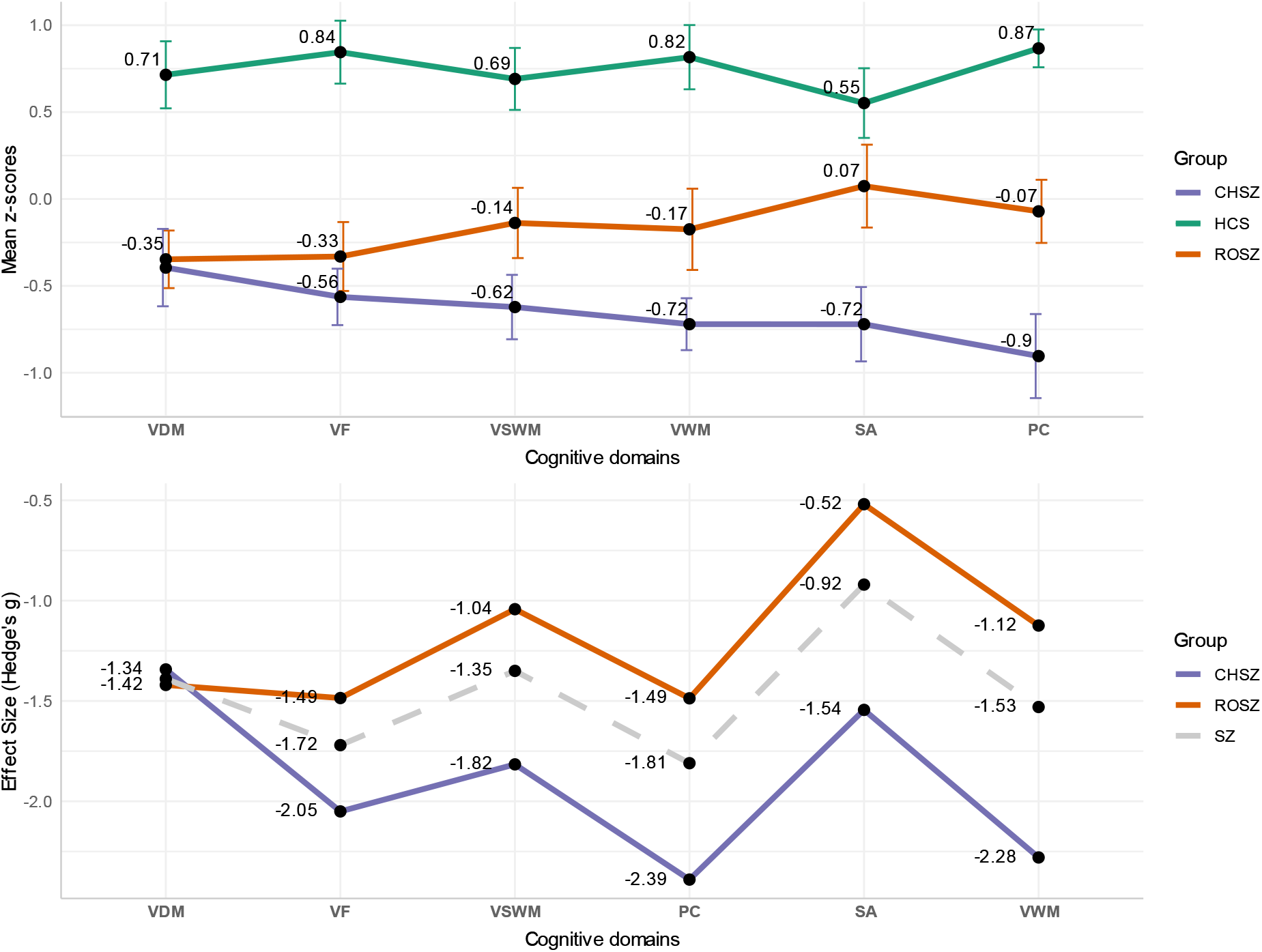
Mean *z*-scores with standard error bars across cognitive domains in HCS, ROSZ and CHSZ (upper panel), and their effect sizes (Hedge’s g) of difference from HCS in ROSZ and CHSZ (lower panel). The domains are arranged in the ascending order of the magnitudes of difference between ROSZ and CHSZ groups. The dashed line in the lower panel is the effect size plotted for both the patient groups combined. See **Tables S1 and S2** in the supplementary material for all effect size estimates with their 95 % confidence intervals.

In ROSZ group, the largest effect size of difference in mean *z*-score units from healthy adults was observed for verbal fluency and perceptual comparison speed (*g* = −1.49). Perceptual comparison speed in the CHSZ sample (*g* = −2.39) had the largest effect size amongst all the six cognitive domains (see **Figure 2**)

### 3.1 Multivariate effects of group on cognitive performance, in the overall sample (*n* = 47)

Multivariate GLM (MANCOVA) revealed significant main effect of group [Pillai’s trace = 0.69, *F*(12, 72) = 3.15, *p* = 0.001] on the six cognitive domain scores taken as dependent variables, after adjusting for mean-centered age, education, and group-by-age interaction (added to account for the possible differential effects of aging, as the three groups differed significantly in age distribution; see **Table 1** and **Figure S3** in supplementary material).We also observed a significant effect of age on GNA performance [Pillai’s trace = 0.30, *F*(6, 35) = 2.47, *p* = 0.042] (see **Table *4***).

In post-hoc univariate analysis (ANCOVA), all the cognitive domains showed significant main effect of group, at an uncorrected threshold of α = 0.05. Perceptual comparison speed, verbal fluency, verbal working memory and verbal declarative memory survived Bonferroni correction for six univariate tests. Perceptual comparison speed had the largest effect size [*F*(2, 40) = 13.92, *p* < 0.001, η^2^_partial_ **=** 0.41] in univariate tests (see **Table 5**).

**Table 5.** Univariate main effects^a^ of group on cognitive domains (post-hoc)

| Cognitive domain | $F(2, 40), p$-value | Bonferroni corrected $p$-value | $\eta^2_{\text{partial}}$ |
| --- | --- | --- | --- |
| Perceptual comparison | 13.92, $p < 0.001^*$ | $< 0.001$ | 0.41 |
| Verbal fluency | 12.18, $p < 0.001^*$ | $< 0.001$ | 0.38 |
| Verbal working memory | 7.63, $p = 0.002^*$ | 0.009 | 0.28 |
| Verbal declarative memory | 7.32, $p = 0.002^*$ | 0.012 | 0.27 |
| Simple attention | 5.04, $p = 0.011$ | 0.067 | 0.20 |
| Visuo-spatial working memory | 4.72, $p = 0.014$ | 0.087 | 0.19 |

<sup>a</sup> Univariate GLM (ANCOVA) adjusted for age, education, and group × age; \* Significant at Bonferroni corrected threshold of $\alpha = 0.05/6 = 0.0083$ ; the cognitive domains are listed in the decreasing order of effect size

### 3.2 Multivariate analysis of neurocognitive performance between healthy (*n* = 16) and schizophrenia samples (*n* = 31)

In multivariate analysis, there was a significant difference between healthy and schizophrenia samples in neurocognitive performance scores [Pillai’s trace = 0.59, *F*(6, 37) = 8.92, *p* < 0.001], after adjusting for mean-centered age, education, and group-by-age interaction (see **Table 6**). Additionally, we observed a significant main effect of age on cognitive scores, although the effect was smaller than the main effect of group [*F*(6, 37) = 2.45, *p* = 0.043, η^2^_partial_ = 0.28].

**Table 6.**
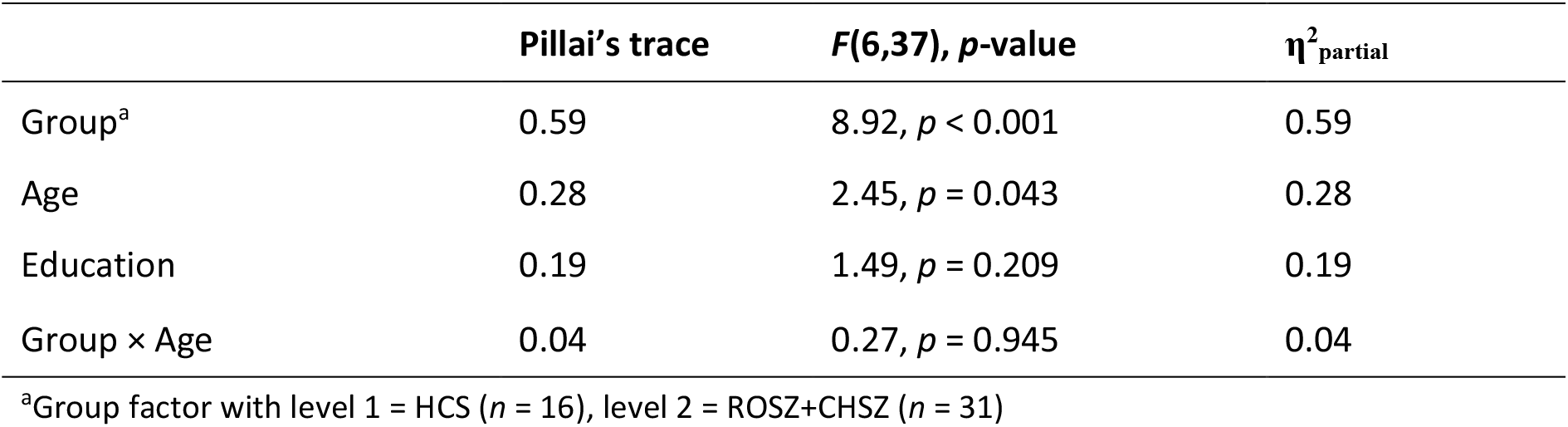
Multivariate GLM (MANCOVA) in healthy versus schizophrenia samples

In post-hoc univariate analysis, all the six domains were significantly different between healthy and schizophrenia groups (*p* < 0.001). All the domains except for simple attention survived Bonferroni correction for six tests (see **Table 7**).

**Table 7.** Univariate main effects (post-hoc) in healthy versus schizophrenia samples

| Cognitive domain | $F(1, 42)$, Bonferroni corrected $p$-value | $\eta^2_{\text{partial}}$ |
| --- | --- | --- |
| Perceptual comparison | 29.35, $p < 0.001^*$ | 0.41 |
| Verbal fluency | 29.11, $p < 0.001^*$ | 0.41 |
| Verbal working memory | 21.21, $p < 0.001^*$ | 0.34 |
| Verbal declarative memory | 16.40, $p = 0.001^*$ | 0.28 |
| Visuo-spatial working memory | 16.38, $p = 0.001^*$ | 0.28 |
| Simple attention | 6.88, $p = 0.072$ | 0.14 |

<sup>a</sup> Univariate GLM (ANCOVA) adjusted for age, education, and group × age; \* Significant at Bonferroni corrected threshold of $\alpha = 0.05/6 = 0.0083$ ; the cognitive domains are listed in the decreasing order of effect size

### 3.3 Comparative analysis of neurocognitive performance between recent-onset (*n* = 17) and chronic schizophrenia (*n* = 14) samples

In the overall schizophrenia sample (recent-onset and chronic combined), we performed a multivariate GLM (MANCOVA) to evaluate the main effect of patient group on the cognitive scores (see **Table 8**). To avoid multicollinearity, age was not included in the model as there were significant between-group differences in age (see **Table 1** and **Figure S3**). Age at onset of symptoms was added as a covariate in view of its link to cognitive impairment (Dorofeikova et al., 2018) and also to account for the potential bias of patients with earlier age at onset being overrepresented in the chronic schizophrenia group. The cumulative dose-years of antipsychotic exposure were substantially different between ROSZ and CHSZ; therefore, we included the average daily dose of antipsychotics (cumulative antipsychotic exposure divided by the total duration of treatment) as a covariate, to avoid multicollinearity. Anticholinergic (trihexyphenidyl) dose was not added in the model due to the negligible difference between ROSZ and CHSZ (see **Table 2**).

**Table 8.** Results of Multivariate GLM (MANCOVA) analysis of neurocognitive performance between recent-onset and chronic schizophrenia samples, controlling for demographic and clinical variables

| | Pillai's trace | $F(6, 19), p\text{-value}$ | $\eta^2_{\text{partial}}$ |
| --- | --- | --- | --- |
| Group<sup>a</sup> | 0.42 | 2.28, $p = 0.079$ | 0.42 |
| Age at onset | 0.36 | 1.79, $p = 0.155$ | 0.36 |
| Education | 0.35 | 1.70, $p = 0.176$ | 0.35 |
| SANS | 0.38 | 1.97, $p = 0.121$ | 0.38 |
| SAPS | 0.59 | 4.63, $p = 0.005$ | 0.59 |
| ADD<sup>b</sup> | 0.26 | 1.09, $p = 0.404$ | 0.26 |

<sup>a</sup>Group factor (level 1 = CHSZ and level 2 = ROSZ); <sup>b</sup>Average daily dose (ADD) of antipsychotic exposure

All ROSZ patients except one had exposure to only second-generation antipsychotics (SGA), most commonly risperidone, followed by olanzapine, aripiprazole, quetiapine and paliperidone. One patient was on treatment with tablet flupenthixol. Thus, the ROSZ group had negligible cumulative exposure to FGA. In the CHSZ group, majority were currently on SGAs, and two were on combination of FGA (tablet flupenthixol, Injection fluphenazine decanoate) and SGA. Although the exposure to FGA was significantly greater in CHSZ than ROSZ (see **Table 2**), the overall average share of FGA in the cumulative antipsychotic exposure of CHSZ group was only 10%. So, our schizophrenia sample consisted of patients predominantly exposed to SGAs.

Analysis of multivariate differences between ROSZ and CHSZ did not reveal significant group effect at *p* < 0.05 threshold, after adjusting for the covariates [*F*(6, 19) = 2.28, *p* = 0.079].

Post-hoc univariate analyses were done to explore the pattern of univariate group effects, keeping in mind the possibility that multivariate main effect of group could have been subtle, and hence not detected in our small sample (see **Table 9**).

**Table 9.** Univariate GLM (ANCOVA) analysis of cognitive domain scores between recent-onset and chronic schizophrenia samples (post-hoc)

| Cognitive domain | $F(1, 24);$<br>uncorrected $p\text{-value}$ | Bonferroni<br>corrected $p\text{-value}$ | $\eta^2_{\text{partial}}$ |
| --- | --- | --- | --- |
| Perceptual comparison | 8.04, $p = 0.009$ | 0.054 | 0.25 |
| Visuo-spatial working memory | 4.70, $p = 0.040$ | 0.240 | 0.16 |
| Simple attention | 3.95, $p = 0.058$ | 0.348 | 0.14 |
| Verbal fluency | 2.14, $p = 0.156$ | 0.936 | 0.08 |
| Verbal working memory | 1.55, $p = 0.224$ | 1.000 | 0.06 |
| Verbal declarative memory | < 0.00, $p = 0.974$ | 1.000 | < 0.00 |

<sup>a</sup> Univariate GLM (ANCOVA) adjusted for age at onset, education, SANS, SAPS, and average daily dose (ADD) of antipsychotics

In univariate analyses, group effects were significant for the PC [*F*(1, 24) = 8.04, *p* = 0.009, η^2^_partial_ = 0.25], and VSWM [*F*(1, 24) = 4.70, *p* = 0.04, η^2^_partial_ = 0.16] cognitive domains, although neither survived Bonferroni correction for multiple univariate tests (corrected threshold α = 0.008). Thus, the overall difference between ROSZ and CHSZ was subtle, with a trend for disproportionately greater impairment in PC domain, which possibly could have been more evident in a larger sample.

### 3.4 Gray matter correlates of the cognitive scores

In the overall sample (*n* = 47) we looked for gray matter volumetric correlates of the cognitive scores in six domains. We found significant positive correlation between perceptual comparison speed scores and gray matter volume of the left anterior-medial temporal lobe and adjacent regions, after adjusting for TIV and age, at a stringent correction for multiple comparisons (TFCE, *p* < 0.0083, FWE corrected). Right anterior medial temporal lobe regions also showed significant positive correlation, albeit to a lesser extent and magnitude (see **Figure 3** and **Table 10**).

**Figure 3.**
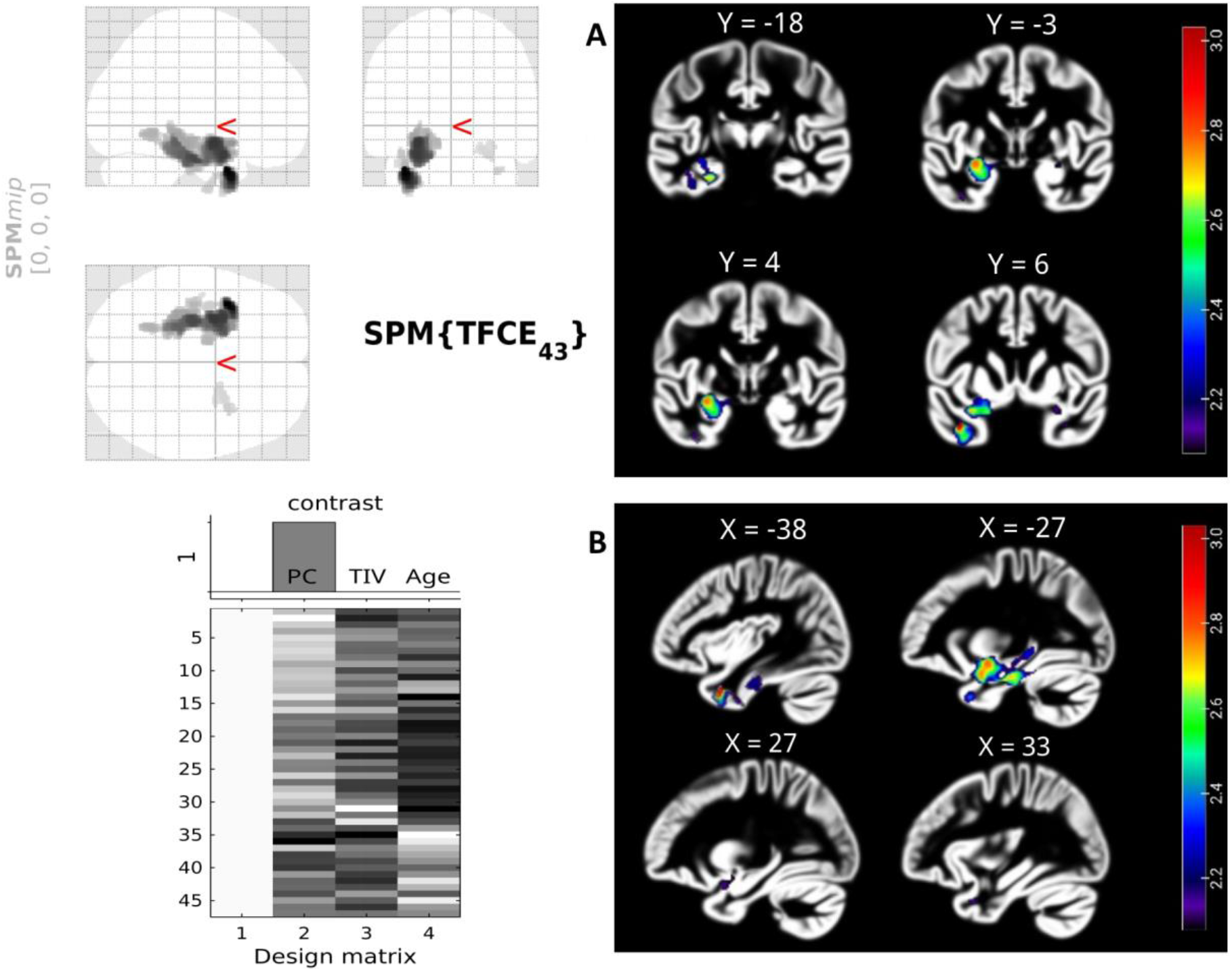
Voxel-wise correlational analysis results showing gray matter clusters with statistically significant positive correlation (TFCE, FWE *p* < 0.0083) with perceptual comparison (PC) speed scores, adjusted for TIV and age (*n* = 47). Coronal and sagittal slices are displayed in neurological convention (left side corresponds to the left hemisphere) and show significant clusters in bilateral anterior and medial temporal lobe regions, predominantly in the left hemisphere (lower row in B shows right hemisphere). The slices were chosen based on the MNI coordinates of the peak voxel in each cluster. The gradient of the colormap is based on negative logarithm of *p*-values with a minimum threshold at 2.0792 (*p*=0.0083).

**Table 10.**
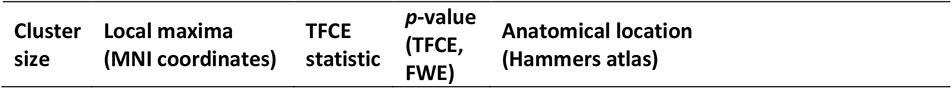
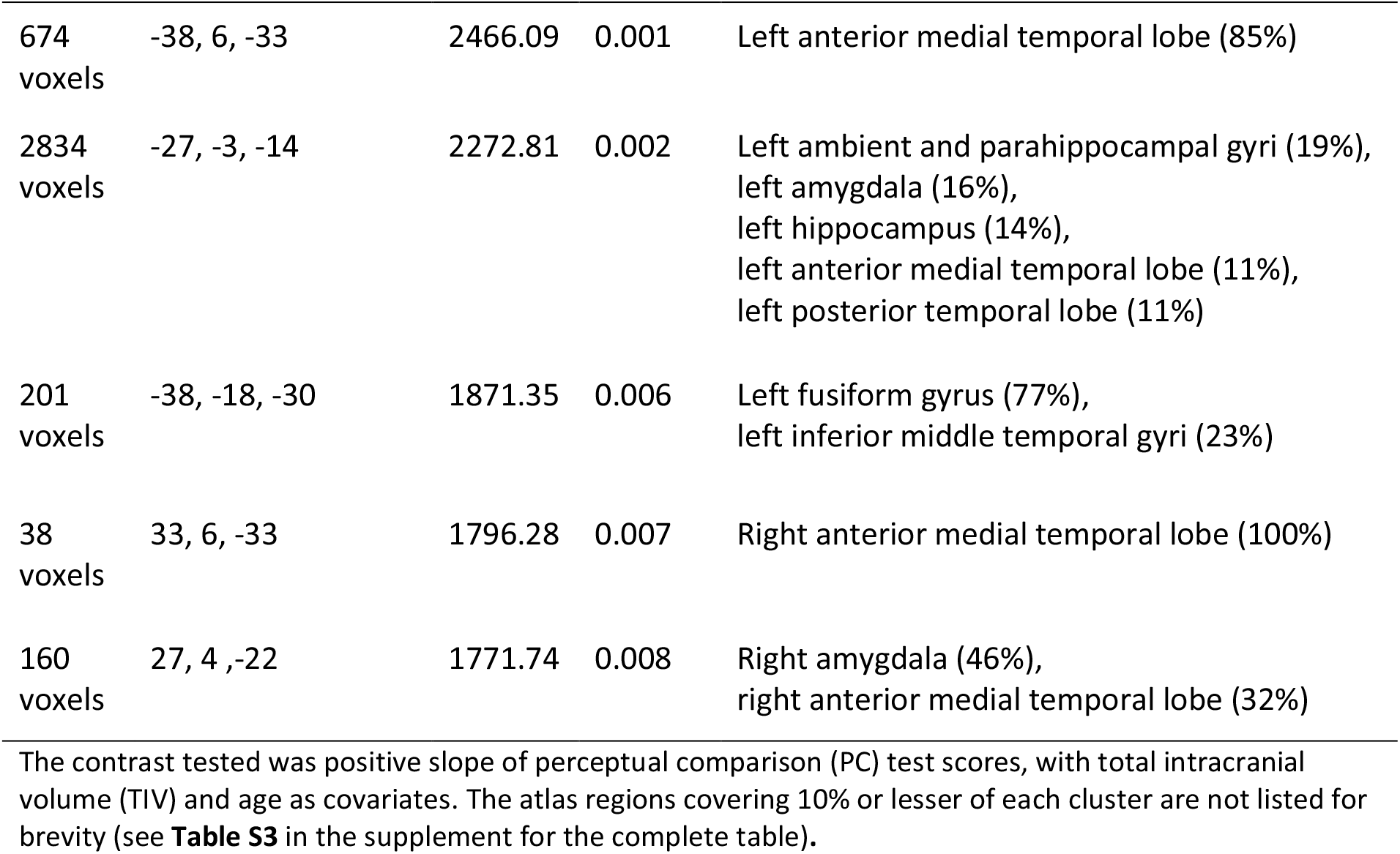
The gray matter clusters with significant correlation with perceptual comparison scores

| Cluster size | Local maxima (MNI coordinates) | TFCE statistic | $p$-value (TFCE, FWE) | Anatomical location (Hammers atlas) |
| --- | --- | --- | --- | --- |
| 674<br>voxels | -38, 6, -33 | 2466.09 | 0.001 | Left anterior medial temporal lobe (85%) |
| 2834<br>voxels | -27, -3, -14 | 2272.81 | 0.002 | Left ambient and parahippocampal gyri (19%),<br>left amygdala (16%),<br>left hippocampus (14%),<br>left anterior medial temporal lobe (11%),<br>left posterior temporal lobe (11%) |
| 201<br>voxels | -38, -18, -30 | 1871.35 | 0.006 | Left fusiform gyrus (77%),<br>left inferior middle temporal gyri (23%) |
| 38<br>voxels | 33, 6, -33 | 1796.28 | 0.007 | Right anterior medial temporal lobe (100%) |
| 160<br>voxels | 27, 4, -22 | 1771.74 | 0.008 | Right amygdala (46%),<br>right anterior medial temporal lobe (32%) |

Verbal Working Memory (digit span backward) scores showed significant positive correlation with the volumes of the right precentral gyrus, the left inferior frontal gyrus, and the left anterior lateral temporal lobe (see **Figure 4** and **Table 11**). Other cognitive domains did not show statistically significant correlation with the gray matter volumes after correction for multiple comparisons.

**Figure 4.**
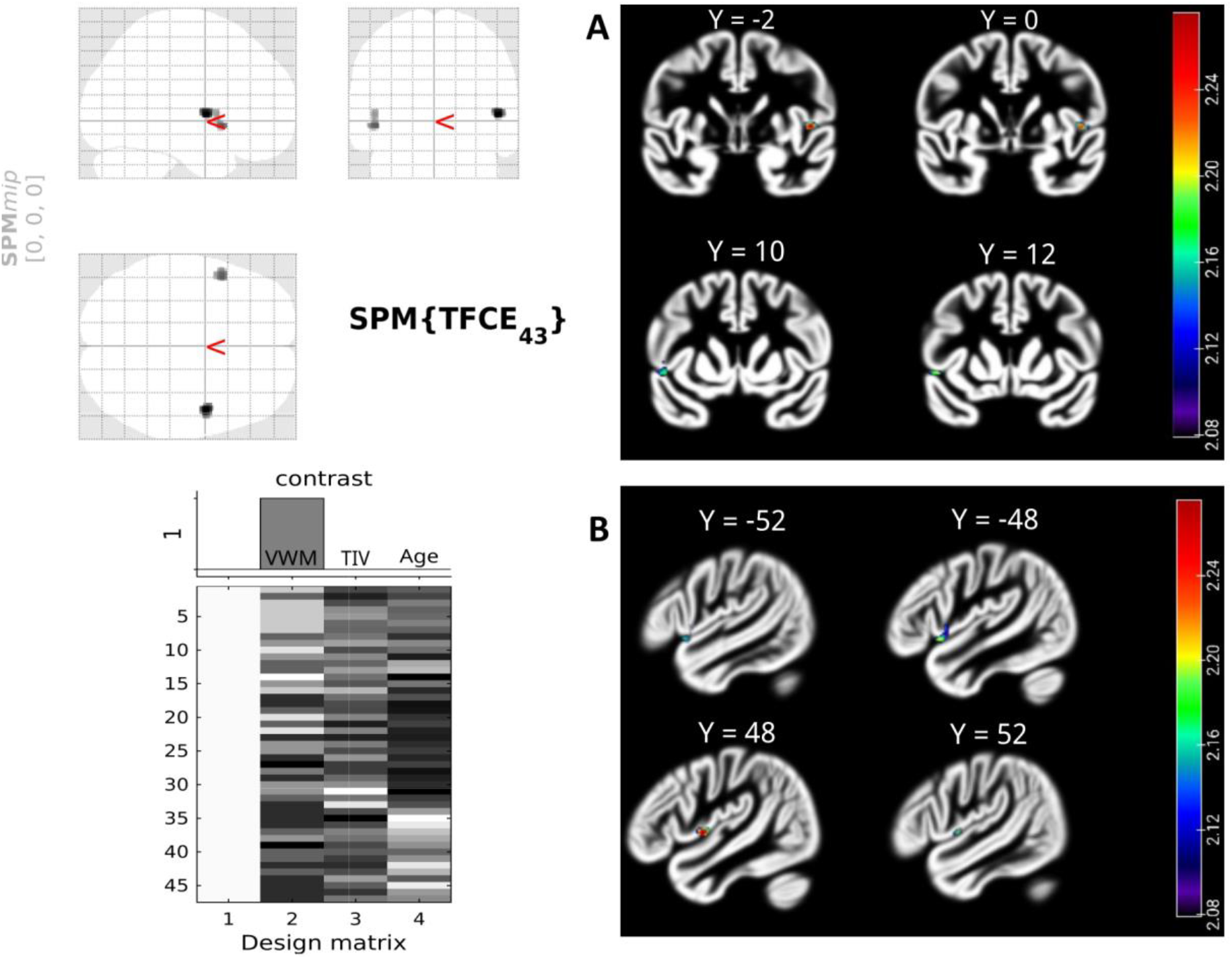
Voxel-wise correlational analysis results showing gray matter clusters with statistically significant positive correlation (TFCE, FWE *p* < 0.01) with verbal working memory (VWM) scores, adjusted for TIV and age (*n* = 47). The coronal and sagittal slices displayed in the neurological convention (left hand side corresponding to the left hemisphere) show significant clusters covering adjacent regions in the precentral gyrus on the right hemisphere (upper row in A and lower row in B), and the inferior frontal gyrus, the anterior lateral temporal lobe, and the precentral gyrus on the left hemisphere (lower row in A and upper row in B). The slices were chosen based on the MNI coordinates of the peak voxel in each cluster. The gradient of the colormap is based on negative logarithm of *p*-values with a minimum threshold at 2.0792 (*p*=0.0083).

**Table 11.** The gray matter clusters with significant correlation with VWM scores

| Cluster size | Local maxima (MNI coordinates) | TFCE statistic | $p$-value (TFCE, FWE) | Anatomical location (Hammers atlas) |
| --- | --- | --- | --- | --- |
| 92 voxels | 48, -2, 3 | 1825.36 | 0.005 | Right precentral gyrus (92%) |
| 91 voxels | -48, 12, -6 | 1768.57 | 0.006 | Left inferior frontal gyrus (56%),<br>left anterior lateral temporal lobe (29%) |

## 4. Discussion

In this pilot study using GNA, we examined the patterns of neurocognitive impairment in recent onset vs. chronic schizophrenia and studied the relationship between these deficits and brain morphometry to explore the link, if any, between cognitive deficits in schizophrenia and brain structural alterations.

### 4.1 Pattern of neurocognitive deficits in schizophrenia— Generalized, selective or both

Using the Global Neuropsychological Assessment, we found an overall pattern of cognitive impairment affecting multiple domains that is consistent with previous reports of generalized cognitive impairment in the schizophrenia literature (Dragovic et al., 2016). All the cognitive domains showed large effect size (|*g*| > 0.8) of difference from the healthy comparison sample, except simple attention in ROSZ group that had a medium effect size (|*g*| ≈ 0.5). This exploratory study also revealed differential impairments across these cognitive domains in recent onset vs. chronic schizophrenia. Perceptual comparison and VF showed the largest effect sizes (*g* = −1.49) in our sample of recent-onset patients. Perceptual comparison showed an even larger effect size in our chronic schizophrenia sample (*g = −* 2.39) (see **Figure 2**). The GNA Perceptual Comparison test is a measure of processing speed (Olson et al., 2021).

Cross-sectional neuropsychological studies in schizophrenia have found disproportionate impairment of processing speed and verbal memory against a background of generalized cognitive dysfunction (Dickinson et al., 2008). Impaired verbal memory and processing speed have therefore been hypothesized to be central deficits that mediate the more generalized cognitive dysfunction in schizophrenia (Andersen et al., 2013). Meta-analyses have also concluded that verbal declarative memory and processing speed show the largest effect size amongst cognitive deficits in first-episode and early schizophrenia (Mesholam-Gately et al., 2009; Zhang et al., 2019).

Verbal fluency (VF) impairment, especially in semantic-category fluency is considered a key neurocognitive feature of schizophrenia, showing large effect sizes in neuropsychological and functional neuroimaging studies (John et al., 2011; Szöke et al., 2008). In our ROSZ sample, VF and PC had the largest effect sizes (*g* = −1.49) amongst the cognitive domains studied. Category VF test performance relies on semantic word generation as well as executive functioning and processing speed (Shao et al., 2014). As VF test is time-limited, one would expect the test performance to positively correlate with processing speed. This was the case in our overall sample, with VF and PC scores showing a strong correlation [*r*(45) = 0.67, *p* < 0.001; **Figure S2**]. Our samples had an age range of 18 – 50 years, a period during which the general intelligence and verbal abilities have been shown to have a trajectory of improvement as part of normal aging, before the age-related decline sets in (Elgamal et al., 2011). The magnitude of difference in the effect sizes of PC (0.90) was substantially larger than that of VF (0.56) between ROSZ and CHSZ groups (see **Figure 2**). This disparity, despite a strong correlation between these two domains, may be because the CHSZ subjects were on an average, older (≈ 41 years) than ROSZ (≈ 24 years) and healthy (≈ 31 years) subjects, and aging might have had a moderating effect through the improvement in lexical semantic knowledge (Elgamal et al., 2011; Salthouse, 2019).

### 4.2 Neurocognitive differences between recent-onset and chronic schizophrenia

In our study, perceptual comparison speed was the chief domain that differed significantly between recent-onset and chronic schizophrenia samples (**Table 9**). Notably, we included ‘age at onset’ rather than ‘age’ as a covariate in ROSZ vs. CHSZ analyses in order to avoid multicollinearity, as age was significantly associated with group, and because of the strong correlation between duration of illness and age, *r*(29) = 0.88, 95% CI [0.76, 0.94]. Furthermore, ‘age at onset’ affects neurocognitive impairment in schizophrenia (Rajji et al., 2009). It is likely that aging also contributed to the slower PC test performance in CHSZ sample, as processing speed is a crucial cognitive function affected by normal aging, and is considered to linearly decline at about 0.02 standard deviation (with reference to young adult norms) per year from the peak at third decade of life until around 80 years of age (Murman, 2015; Salthouse, 2010, 2019).

Our three-group multivariate analysis also showed significant effects of age, though with a smaller effect size than group (**Table 4**). Here we did not attempt post-hoc pairwise group comparisons using age-adjusted means of cognitive scores because of the significant difference in mean ages between the three groups (see **Table 1** and **Figure S3**), a scenario where the estimated marginal means would not be meaningful (Clason and Mundfrom, 2012). Moreover, it is unlikely that older age accounts for the slow processing speed shown by patients with chronic schizophrenia because patients with recent-onset schizophrenia also showed the largest decrement in this domain, and they were younger than the healthy controls. Also, the median age of CHSZ sample was ≈ 40 years, which is too early to observe substantial age-associated cognitive deterioration. Finally, our univariate analysis of PC speed revealed a significant difference between HCS and CHSZ groups after adjusting for age, education, and group-by-age interaction [*F* (1, 25) = 17.53, *p* = 0.0003, η^2^_partial_ = 0.41].

In short, this study provides evidence of disproportionate impairment of perceptual comparison/processing speed in chronic schizophrenia vs. recent-onset schizophrenia. An ideal assessment of the effect of chronicity requires a longitudinal study design; however, it would be cumbersome to model the multitude of variables that contribute to changes in cognitive performance over decades, and also control for practice effects (Goldberg et al., 2010) due to repeated assessments.

Previous cross-sectional studies that have compared recent-onset or first-episode vs. chronic schizophrenia are sparse. A few have found comparable levels of cognitive impairment overall in both groups of patients, with greater impairment of processing speed, working memory and social cognition in chronic compared to recent-onset or first-episode schizophrenia (Liu et al., 2019; McCleery et al., 2014; Sponheim et al., 2010). The comparable magnitude of cognitive deficits in chronic vs. early schizophrenia implies a neurodevelopmental pathology of brain. However, this does not preclude the possibility of an added neuroprogression component. Longitudinal neuroimaging and neuropsychological studies have also found evidence of progression in brain structural changes and cognitive dysfunction in schizophrenia (Andreasen et al., 2011; Olabi et al., 2011). In their review of longitudinal neuroimaging studies (follow-up duration 5 to 21 years), Hulshoff and colleagues concluded that schizophrenia is associated with progressive brain morphometric changes especially in the frontal lobe, and neuroprogression might not be restricted to the early phase of illness (Hulshoff Pol and Kahn, 2008). A recent meta-analysis of longitudinal clinical, cognitive and neuroimaging changes in schizophrenia observed that follow-up intervals longer than 10 years are rare, thus limiting conclusions about the long-term trajectory of these changes (Heilbronner et al., 2016). The few studies that followed up patients for 15-20 years report overall stability of cognitive deficits over time until around 50 years of age, with probable accelerated decline in executive functions in later life compared to healthy controls, albeit of small effect size (Albus et al., 2020; Bonner-Jackson et al., 2010; Herold et al., 2017; Irani et al., 2012).

There is emerging evidence from neurocognitive, neuroimaging, and metabolic studies for ‘accelerated aging’ in schizophrenia (Kirkpatrick and Kennedy, 2018; Schnack et al., 2016). The neuropsychological premise of this hypothesis is that the cognitive profile of patients with schizophrenia is similar to that of older healthy adults, with verbal memory, executive functions, and processing speed as the main affected domains (Dragovic et al., 2016). The profile of impairment observed in the overall schizophrenia sample of our study also agrees with this pattern (see **Table 7**). It may be noted that other than VDM, all the five domains showed substantial differences between the ROSZ and CHSZ groups, with effect sizes ranging from 0.56 for VF to 1.16 for VWM (see **Figure 2**). However, after adjusting for the effects of symptoms, age at onset, education and daily antipsychotic dose, PC showed the largest difference between ROSZ and CHSZ (η^2^_partial_ = 0.25), while VWM (η^2^ _partial_ = 0.06) and VDM (η^2^_partial_ ∼ 0.00) showed small to negligible differences (see **Table 9**). This is consistent with previous longitudinal studies that have reported more pronounced age-related decline in processing speed compared to verbal memory (Lee et al., 2020). In normal aging, processing speed starts declining early in the middle age (Salthouse, 2019; Weaver Cargin et al., 2007; Zaninotto et al., 2018), whereas verbal declarative memory tends to begin declining after about age 60 (Brickman and Stern, 2009). Around age of ≈ 40 years (median age of our CHSZ sample), one would not expect a marked decline in verbal declarative memory as part of normal aging (Harada et al., 2013). It is also pertinent to note that early cognitive changes in classical neurodegenerative disorders like Alzheimer’s Disease are characterized by significant deficits in verbal learning/memory and processing speed domains that are comparable in magnitude (Bastin and Salmon, 2014; Constantinidou et al., 2014; Lange et al., 2002; Tabert et al., 2006). Thus, the picture of a disproportionate deficit in processing speed and negligible deficit in verbal declarative memory in our CHSZ sample could indicate a pattern of accelerated or premature cognitive aging, unlike the pattern seen in classical neurodegeneration.

Attention and working memory (WM) are closely associated cognitive functions. Digit span forward (DSF) tests simple attention. Digit span backward (DSB) requires greater cognitive control and places greater demands on the WM system (Donolato et al., 2017; St Clair-Thompson and Allen, 2013). VSWM was tested by spatial span forward and backward tests in the GNA. In our results, DSF and DSB tests in the GNA showed strong correlation [*r*(45) = 0.72, *p* < 0.001], both tests being measures of verbal short-term retention; whereas VSWM showed moderate correlation with verbal WM [*r*(45) = 0.49, *p* < 0.001], probably because of the different neural processes underlying these WM sub-domains — the visuospatial sketchpad and phonological loop respectively (Baddeley, 2012). VSWM showed a stronger correlation with PC speed domain [*r*(45) = 0.69, *p* < 0.001] (See **Figure S2**), possibly because both tests entail visual processing. Meta-analyses have concluded that WM dysfunction is a consistent feature of schizophrenia, and the magnitude of this dysfunction does not differ significantly across verbal and visuospatial WM tasks (Forbes et al., 2009; Lee and Park, 2005). Forbes et al. further reported that WM performance was significantly worse in chronic/multiple-episode schizophrenia compared to first-episode patients, even though WM scores did not correlate with duration of illness. They argued that this might reflect accelerated cognitive aging (Forbes et al., 2009). In our study, both verbal and visuospatial WM tasks revealed impairment in the schizophrenia sample, while the deficit in simple attention was less marked (see **Table 7**). Overall, the verbal and visuospatial domains of WM were both impaired in CHSZ and ROSZ samples to a similar extent, in comparison to healthy sample (see **Figure 2**). This is consistent with the pattern of deficits in WM and in its sub-domains reported in schizophrenia literature (Park and Gooding, 2014).

### 4.3 The neuroanatomical correlates of cognitive deficits in schizophrenia

In our voxel-wise correlational analysis, perceptual comparison/processing speed performance correlated positively with gray matter volume in anterior and medial temporal lobe regions, predominantly of left side (see **Figure 3** and **Table 10**). The GNA Perceptual Comparison test requires one to make ‘same/different’ judgments about as many pairs of simple geometric shapes as possible in 45 seconds (see **Figure S1** in supplementary material for details). Processing speed is a complex construct employing diverse neural networks depending on the type of stimulus being processed (Wong et al., 2021). Thus, the GNA PC test requires speeded visual scanning, processing of geometric shapes, and oral responses that minimize motor system demands imposed by commonly used paper-and-pencil tests of processing speed, such as the Trail Making Test and Digit Symbol Substitution Test.

The anterior and medial temporal lobe regions have been consistently implicated in schizophrenia pathogenesis and its clinical presentation (Bobilev et al., 2020; DeRamus et al., 2020; Kaur et al., 2020). The anterior temporal lobe (ATL) is crucial for verbal and non-verbal semantic processing (Binney et al., 2010; Snowden et al., 2018), and is considered a ‘transmodal hub’ (the ‘hub-and-spoke’ hypothesis of semantic memory) involved in processing semantic aspects of stimuli across sensory modalities (Lambon Ralph et al., 2010). There is also increasing evidence for ‘gradients’ of specificity within ATL, with the anterior and ventromedial regions showing greater specificity for complex visual processing (Bonner and Price, 2013; Patterson and Lambon Ralph, 2016). Low frequency repetitive transcranial magnetic stimulation to the ATL, especially the left side, can significantly slow reaction times in visual semantic processing tasks that also involve speech production, like picture naming (Pobric et al., 2007; Woollams et al., 2017).

Recent evidence supports the key role of medial temporal lobe (MTL) and adjacent structures – the hippocampus, entorhinal cortex, perirhinal cortex and parahippocampal cortex, not just for memory, but also for visual perceptual processing (Moscovitch et al., 2016). Neuropsychological studies in patients with MTL damage, and functional MRI studies in healthy individuals provide strong evidence for the involvement of hippocampus and perirhinal cortex in visual discrimination tasks, especially when there are similarities and shared features between the simultaneously presented stimuli (Bonnen et al., 2021; Lawrence et al., 2020; Lech and Suchan, 2014). Because the GNA PC test entails comparing pairs of geometric shapes that are presented simultaneously, it does not require visual memory. Given the evidence supporting the role of anterior and medial temporal lobes in visual perceptual comparison, it is reasonable to conclude from the findings of our study that subtle deficits in these brain regions may underlie the impairment in perceptual comparison speed seen in schizophrenia.

We found small clusters in the right precentral gyrus, left inferior frontal gyrus, left anterolateral temporal lobe, and left precentral gyrus showing significant positive correlation with verbal working memory scores (**Figure 4**). The brain areas underlying verbal working memory are diverse, as multiple processes like speech perception, temporary storage, manipulation, and speech output are involved (Shura et al., 2016). The left inferior frontal gyrus is considered a key area for subvocal rehearsal in Baddeley’s phonological loop model (Baddeley, 2003). The left perisylvian areas have been closely linked to digit repetition, especially backward digit span, in healthy adults (Zhou et al., 2006) and patients with lesions involving this region (Geva et al., 2021; Koenigs et al., 2011; Papagno et al., 2017). The inferior precentral gyrus, the superior temporal gyrus, and the insula are structurally and functionally interconnected areas (Ghaziri et al., 2017), and have been implicated in working memory functioning (Miotto et al., 2014; Tomasino and Gremese, 2016). The inferior precentral gyrus has been shown to be important for short term retention of auditory stimuli before further processing and motor transformation (Carpenter et al., 2018; Kambara et al., 2018; Nakai et al., 2017). Structural and functional aberrations in the perisylvian cortical regions like the precentral gyrus, the inferior frontal gyrus, and the superior temporal gyrus are widely implicated in the auditory processing dysfunction that may underlie auditory hallucinations in schizophrenia (Javitt and Sweet, 2015; John et al., 2017). Overall, our findings are in line with the existing literature on neural correlates of phonological working memory; however, these preliminary results with modest effect sizes need replication in larger samples of patients with recent-onset and chronic schizophrenia.

### 4.4 The effects of symptoms and medications on cognitive performance

There is considerable evidence that negative symptoms have direct association with cognitive dysfunction (Marder and Galderisi, 2017; Strassnig et al., 2018), while positive symptoms have a distracting effect on cognitive task performance (Moritz et al., 2021). Additionally, greater degree of positive symptoms warrants higher dose of medications like antipsychotics, anticholinergics, benzodiazepines etc. which can impede cognitive performance through anticholinergic effects (Ang et al., 2017; Frydecka et al., 2016), sedation, and psychomotor slowing due to extrapyramidal effects (Caligiuri et al., 2019; Knowles et al., 2010). In our study, we excluded patients who were on benzodiazepines to prevent some of these confounding factors. The ROSZ group in our study had higher cross-sectional positive and negative symptom scores than CHSZ, and the difference was statistically significant for PSYRATS scores (see **Table 2**). SAPS scores significantly contributed to reduction in cognitive scores in our schizophrenia sample with small-moderate effect size, as shown in our multivariate analysis between ROSZ and CHSZ that adjusted for average daily antipsychotic dose as well as age at onset, education, and SANS scores (**Table 8**). Our sample consisted of patients who were cooperative and motivated for various assessments and subsequent MR imaging, and therefore did not include patients with prominent negative symptoms. This unavoidable selection bias, along with the small sample size, might explain why we did not find significant effects of SANS ratings on cognitive performance.

Multiple meta-analyses have reported that long-term antipsychotic exposure is associated with reduction in cortical gray matter (GM) volumes (Fusar-Poli et al., 2013; Vita et al., 2015). However, there could be differential effects of antipsychotics based on their mechanisms of action (FGA vs SGA), and the regions of brain affected (different lobes, cortical vs sub-cortical). There is some evidence, though inconsistent, for a differential cortical GM loss associated with FGAs in comparison to SGAs (Lieberman et al., 2005; Vita et al., 2015). There is also evidence for increase in sub-cortical GM volume especially of basal ganglia, attributed more to FGAs than SGAs (Huhtaniska et al., 2017). Although earlier studies had reported SGAs to be superior to FGAs in improving cognitive deficits in schizophrenia, more recent studies provide conflicting evidence (Davidson et al., 2009; Husa et al., 2017; Keefe et al., 2007; Veselinović et al., 2019). Given the myriad medication associated factors that might impact cognitive functioning, like anticholinergic effects, extrapyramidal effects, metabolic effects, and varying neuronal receptor affinities within and between antipsychotic classes, it would be premature to draw conclusions regarding selective advantages or disadvantages of a particular agent or class of antipsychotics. Exposure to FGAs in our samples was minimal, accounting for less than 1 % of the cumulative antipsychotic exposure in ROSZ group, and 10 % in CHSZ group. The ongoing daily antipsychotic dose was significantly higher in the CHSZ group (see **Table 2**); a similar trend has been noted in previous studies as well (Emsley et al., 2013). However, the daily anticholinergic doses and extrapyramidal side effects were not significantly different between the groups (see **Table 2**). Even with substantially greater cumulative medication exposure, the BMI of CHSZ sample was not significantly higher than ROSZ sample (**Table 1**). Therefore, we speculate that the differential cognitive deficits observed in our ROSZ and CHSZ samples are less likely due to anticholinergic dose differences or the extrapyramidal and metabolic side effects of antipsychotics.

### 4.5 The utility of GNA as a brief cognitive assessment battery in schizophrenia

This is the first study to demonstrate the usefulness of the GNA to detect both generalized and differential cognitive impairments in adults with recent-onset or chronic schizophrenia. The brevity of GNA confers the advantage of limiting the effect of fatigue on cognitive performance, which is especially important in research on patients with schizophrenia (Irani et al., 2012). There is only minimal influence of language and culture on the test items, and literacy is not a pre-requisite. GNA covers most of the cognitive domains affected in schizophrenia that are included in the MATRICS consensus cognitive battery (MCCB) (Nuechterlein et al., 2008), except for reasoning/problem solving and social cognition. The process of validation and developing norms of GNA are currently undergoing worldwide (https://gninc.org/home/about/). An internationally accepted, culture- and language-neutral, brief neuropsychological battery could prove to be immensely useful in the clinical and research settings to study a widely prevalent illness like schizophrenia.

## 5. Conclusions

Patients with recent-onset and chronic schizophrenia had comparable magnitudes of cognitive deficits affecting multiple cognitive domains, with a trend for selectively greater impairment in perceptual comparison/processing speed in the chronic group. This, along with only negligible difference in verbal declarative memory performance between these groups might reflect a pattern indicating accelerated or premature cognitive aging in schizophrenia.

Further, volumetric deficit in the left anterior-medial temporal lobe may underlie the perceptual comparison speed deficit seen in schizophrenia, which if replicated using larger samples can be considered a target for transcranial neuromodulation interventions to improve processing speed.

## Supporting information

STROBE checklist

## Data Availability

All data produced in the present study are available upon reasonable request to the authors

## 6. Author contributions

**Vineeth Mohan:** Data curation, Methodology, Writing-Original draft preparation, Investigation, Formal analysis. **Pravesh Parekh:** Data curation, Writing-Review and Editing, Investigation, Software, Visualization. **Ammu Lukose:** Investigation, Writing-Review and Editing. **Sydney Moirangthem:** Supervision, Fund acquisition, Writing-Reviewing and Editing. **Jitender Saini:** Supervision, Fund acquisition, Writing-Reviewing and Editing. **David J Schretlen:** Development of the GNA, Writing-Review and Editing. **John P John:** Conceptualization, Writing-Review and Editing, Funding acquisition, Resources, Supervision.

## 7. Acknowledgements

We acknowledge the contributions of Mr Krishnendu Vyas, Junior Research Fellow, Multimodal Brain Image Analysis Laboratory (MBIAL) and Mr Abhineet Ojha, Senior Research Fellow, MBIAL in data acquisition. We also acknowledge Ms Rashmi Rao, Mr Anand, Mr Bopanna, and Ms Gomathi at the ADBS Neuroimaging Centre (ANC) for their assistance in MRI acquisition.

## 8. Funding

VM acknowledges funding support from Indian Council of Medical Research (ICMR) under Talent Search Scheme (TSS). PP acknowledges salary support from the Department of Biotechnology (DBT), Government of India (BT/PR17316/MED/31/326/2015). The study utilized funding support from Department of Science and Technology (DST), Government of India to JPJ (DST/INT/TUNISIA/P-11/2017) for MRI acquisition.

## 9. Conference presentation

Part of the findings of this study was presented as an e-poster at the ninth International Conference on Schizophrenia (IConS) of Schizophrenia Research Foundation (SCARF) on July 24^th^ to 31^st^, 2021.

## 10. Declarations of interest

The authors had no competing interests.

## Notes

### Competing Interest Statement

The authors have declared no competing interest.

### Author Declarations

Ethics committee of National Institute of Mental Health and Neurosciences gave ethical approval for this work [NO.NIMH/DO/IEC (BEH.Sc.DIV)/2019, dated 25th May 2019]

