## Supplementary material for "Patterns of impaired neurocognitive performance on Global Neuropsychological Assessment (GNA), and their brain structural correlates in recent-onset and chronic schizophrenia: A pilot study"

**Perceptual comparison test in Global Neuropsychological Assessment (GNA) battery**

In the perceptual comparison (PC) test, the examinee is asked to answer whether the sets of geometric shapes given on the two sides of an item are the ‘same’ or ‘different’. The test consists of 54 such items arranged in three columns. The total number of correctly answered items in 45 seconds are summed as the raw score.

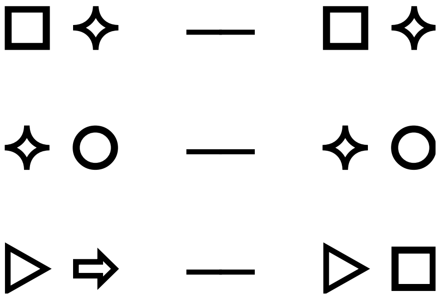

**Figure S1**. Sample of the perceptual comparison test in GNA, showing three items. The correct responses for these are “same,” “same,” and “different.”

### <u>Correlation between cognitive domains in the overall sample (*n* = 47)</u>

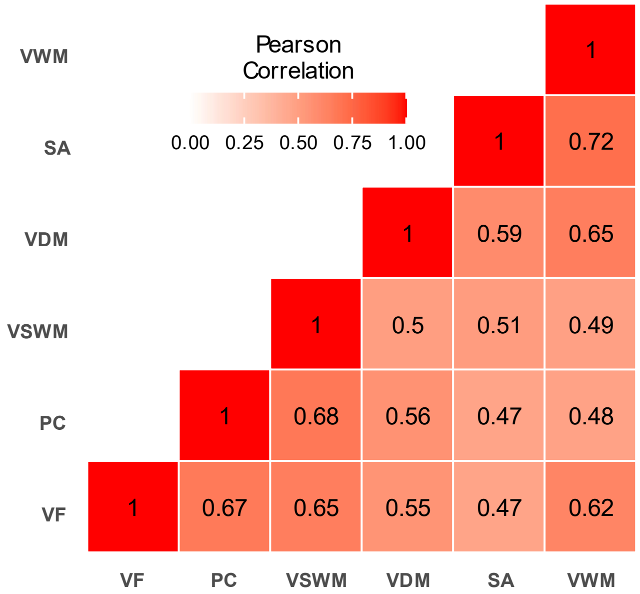


**Figure S2**. Correlation heatmap of the cognitive domains. All the pairwise correlations (Pearson's *r*) were statistically significant at the Bonferroni corrected threshold of $\alpha_b$ = 0.05/15 = 0.0033; VF, Verbal fluency; PC, Perceptual comparison; VSWM, Visuospatial working memory; VDM, Verbal declarative memory; SA, Simple attention; VWM, Verbal working memory.

### Age distribution in the three groups

The age distribution of the three groups differed significantly in the order CHSZ > HCS > ROSZ. Due to the poor age overlap between the recent-onset and the chronic schizophrenia samples, a direct comparison between these patient groups adjusting for age as a covariate would be inappropriate due to potential multicollinearity. In such a situation where the covariate is significantly different between groups, the estimated marginal means calculation to find the pairwise differences might not be meaningful (see *Clason, D.L., Mundfrom, D.J., 2012. Adjusted Means in Analysis of Covariance: Are They Meaningful?*).

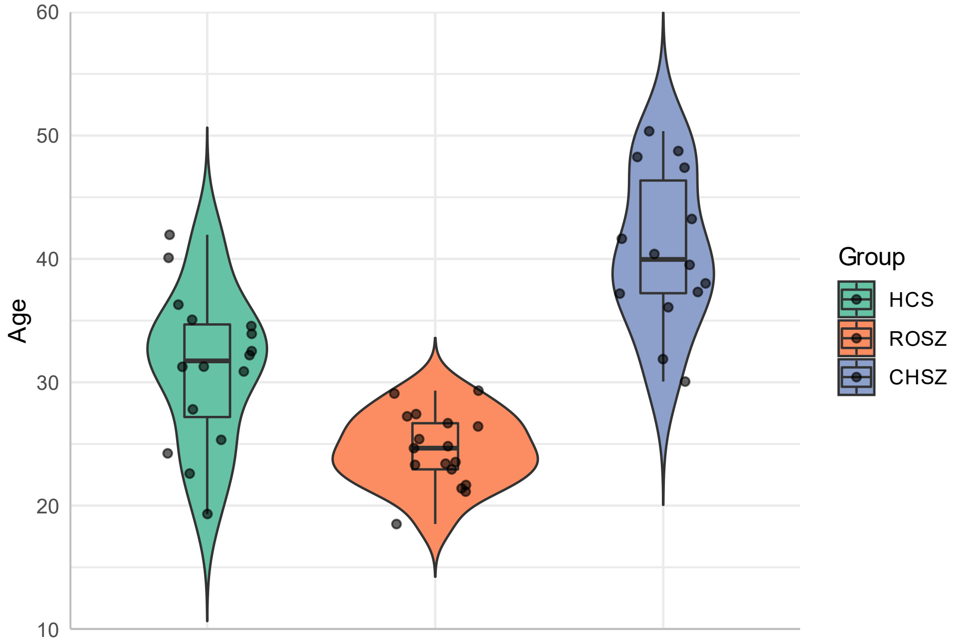


**Figure S3.** Violin plots showing the age distributions in the three groups. As the violin plots are based on kernel density estimates, they extend beyond the range of actual values in the sample depicted by the jittered dots.

### Effect size estimates using different methods for each domain

Effect sizes were estimated assuming unequal variance between groups. We primarily used Hedge's *g* which is less affected by sample size bias than Cohen's *d*, while Glass's Δ uses the standard deviation of the control group (see *Lakens, D., 2013. Calculating and reporting effect sizes to facilitate cumulative science: a practical primer for t-tests and ANOVAs. Front. Psychol*., and *Delacre, M, et al., 2021. Why Hedges' g*s based on the non-pooled standard deviation should be reported with Welch's t-test*)

**Table S1.** Effect sizes of difference in mean cognitive scores between ROSZ and HCS, estimated using different measures, assuming unequal variance between groups

| Domain | Cohen's *d* [95% CI] | Hedge's *g* [95% CI] | Glass's Δ [95% CI] |
|---|---|---|---|
| PC | -1.53 [-2.31, -0.73] | -1.49 [-2.25, -0.71] | -2.16 [-3.36, -0.91] |
| VF | -1.52 [-2.29, -0.73] | -1.49 [-2.24, -0.72] | -1.62 [-2.53, -0.68] |
| VDM | -1.46 [-2.22, -0.67] | -1.42 [-2.17, -0.65] | -1.38 [-2.17, -0.55] |
| VWM | -1.15 [-1.88, -0.41] | -1.12 [-1.84, -0.40] | -1.34 [-2.25, -0.40] |
| VSWM | -1.07 [-1.79, -0.33] | -1.04 [-1.75, -0.32] | -1.16 [-2.00, -0.30] |
| SA | -0.53 [-1.22, 0.17] | -0.52 [-1.19, 0.16] | -0.60 [-1.38, 0.20] |

PC, Perceptual comparison; VF, verbal fluency; VDM, Verbal declarative memory; VWM, Verbal working memory; VSWM, Visuospatial working memory; SA, Simple attention

**Table S2.** Effect sizes of difference in mean cognitive scores between CHSZ and HCS, estimated using different measures, assuming unequal variance between groups

| Domain | Cohen's *d* [95% CI] | Hedge's *g* [95% CI] | Glass's Δ [95% CI] |
|---|---|---|---|
| PC | -2.49 [-3.57, -1.39] | -2.39 [-3.42, -1.33] | -4.07 [-5.92, -2.17] |
| VWM | -2.34 [-3.27, -1.39] | -2.28 [-3.18, -1.35] | -2.08 [-3.03, -1.09] |
| VF | -2.11 [-3.00, -1.20] | -2.05 [-2.91, -1.16] | -1.94 [-2.88, -0.98] |
| VSWM | -1.87 [-2.72, -0.99] | -1.82 [-2.65, -0.96] | -1.84 [-2.79, -0.86] |
| SA | -1.59 [-2.41, -0.75] | -1.54 [-2.34, -0.73] | -1.59 [-2.48, -0.66] |
| VDM | -1.38 [-2.18, -0.56] | -1.34 [-2.12, -0.55] | -1.44 [-2.33, -0.52] |

### Voxel-wise correlation results (SPM 12, v7771) for perceptual comparison speed (PC)

**Table S3.** CAT (version 1727) lookup table using the Hammers atlas, for perceptual comparison (PC) domain

| Cluster size (voxels) | Local maxima (MNI coordinates) | TFCE | $p_{FWE\text{-}corr}$ | Overlap of region (Hammers atlas) | Anatomical location |
|---|---|---|---|---|---|
| 674 | -38, 6, -33 | 2466.09 | 0.001 | 85% | Left anterior medial temporal lobe |
| | | | | 8% | Left fusiform gyrus |
| | | | | 4% | Left inferior middle temporal gyri |
| | | | | 3% | Left anterior lateral temporal lobe |
| 2834 | -27, -3, -14 | 2272.81 | 0.002 | 19% | Left ambient and parahippocampal gyri |
| | | | | 16% | Left amygdala |
| | | | | 14% | Left hippocampus |
| | | | | 11% | Left anterior medial temporal lobe |
| | | | | 11% | Left posterior temporal lobe |
| | | | | 8% | Left insula |
| | | | | 8% | Left orbito-frontal gyri |
| | | | | 5% | Background |
| | | | | 5% | Left lingual gyrus |
| | | | | 2% | Left putamen |
| | | | | 1% | Left cerebellum |
| | | | | 1% | Left lateral temporal ventricle |
| 201 | -38, -18, -30 | 1871.35 | 0.006 | 77% | Left fusiform gyrus |
| | | | | 23% | Left inferior middle temporal gyri |
| 38 | 33, 6, -33 | 1796.28 | 0.007 | 100% | Right anterior medial temporal lobe |
| 160 | 27, 4 ,-22 | 1771.74 | 0.008 | 46% | Right amygdala |
| | | | | 32% | Right anterior medial temporal lobe |
| | | | | 11% | Background |
| | | | | 6% | Right orbito-frontal gyri |
| | | | | 4% | Right insula |

**Voxel-wise correlation results (SPM 12, ver 7771) for verbal working memory (VWM)**

**Table S4.** CAT (version 1727) lookup table using the Hammers atlas, for verbal working memory (VWM) domain

| Cluster size (voxels) | Local maxima (MNI coordinates) | TFCE | $p_{FWE\text{-}corr}$ | Overlap of region (Hammers atlas) | Anatomical location |
|---|---|---|---|---|---|
| 92 | 48, -2, 3 | 1825.36 | 0.005 | 92% | Right precentral gyrus |
| | | | | 8% | Right postcentral gyrus |
| 91 | -48, 12, -6 | 1768.57 | 0.006 | 56% | Left inferior frontal gyrus |
| | | | | 29% | Left anterior lateral temporal lobe |
| | | | | 9% | Left precentral gyrus |
| | | | | 7% | Background |
